# Second wave mortality among patients hospitalised for COVID-19 in Sweden: a nationwide observational cohort study

**DOI:** 10.1101/2021.03.29.21254557

**Authors:** Kristoffer Strålin, Erik Wahlström, Sten Walther, Anna M Bennet-Bark, Mona Heurgren, Thomas Lindén, Johanna Holm, Håkan Hanberger

**Affiliations:** Department of Infectious Diseases, Karolinska University Hospital, Stockholm, Sweden; Department of Medicine, Huddinge, Karolinska Institutet, Stockholm, Sweden; National Programme Area for Infectious Diseases, National System for Knowledge-Driven Management within Healthcare, Sweden’s Regions in Collaboration, Sweden; National Board of Health and Welfare, Sweden; Swedish Intensive Care Registry, Värmland County Council, Karlstad, Sweden; Department of Cardiothoracic and Vascular Surgery, Heart Centre, Linköping University Hospital, Linköping, Sweden; Department of Health, Medicine, and Caring Sciences, Linköping University, Linköping, Sweden; Division of Inflammation and Infection, Department of Biomedical and Clinical Sciences, Faculty of Medicine and Health Sciences, Linköping University, Linköping, Sweden; Department of Infectious Diseases, Linköping University Hospital, Linköping, Sweden

## Abstract

**Background:** During the first pandemic wave, a substantial decline in mortality was seen among hospitalised COVID-19 patients. We aimed to study if the decreased mortality continued during the second wave, using data compiled by the Swedish National Board of Health and Welfare.

**Method:** Retrospective nationwide observational study of all patients hospitalised in Sweden between March 1^st^ and December 31^st^, 2020, with SARS-CoV-2 RNA positivity 14 days before to 5 days after admission and a discharge code for COVID-19. Outcome was 60-day all-cause mortality. Poisson regression was used to estimate the relative risk (RR) for death by month of admission, adjusting for age, sex, socio-economic data, comorbidity, care dependency, and country of birth.

**Findings:** A total of 32 452 patients were included. December had the highest number of admissions/month (n=8253) followed by April (n=6430). The 60-day crude mortality decreased from 24·7% (95% CI, 23·0%-26·5%) for March to 10·4% (95% CI, 8·9%-12·1%) for July-September (as reported previously), later increased to 19·9% (95% CI, 19·1-20·8) for December. RR for 60-day death for December (reference) was higher than those for June to November (RR ranging from 0·74 to 0·89; 95% CI <1 for all months). SARS-CoV-2 variants of concern were only sporadically found in Sweden before January 2021.

**Interpretation:** The decreased mortality of hospitalised COVID-19 patients after the first wave turned and increased during the second wave. Focused research is urgent to describe if this increase was caused by a high load of patients, management and treatment, viral properties, or other factors.

**Research in context:** *Evidence before this study:* During the first pandemic wave, a substantial decline in mortality was seen among hospitalised COVID-19 patients in many countries. As the reason for this decline has not been clarified, no one could foresee how mortality would change during forthcoming waves.

*Added value:* This retrospective nationwide study of all patients hospitalised for COVID-19 in Sweden from March to December 2020 showed that the gradual decrease in mortality seen in the first pandemic wave was followed by an increased crude and adjusted 60-day all-cause mortality during the second wave. This increase in mortality occurred although the standard-of-care recommendations for hospitalised COVID-19 patients did not change in Sweden during the second half of 2020.

*Implications of all the available evidence:* While improved standard-of-care was believed to be an important factor for the decrease in mortality during the first pandemic wave, the increasing mortality during the second wave has no apparent explanation. As the currently known virus variants of concern occurred only sporadically in Sweden before January 2021, they were most likely not involved. Focused research is urgent to describe if this increase in mortality was caused by a high load of patients, management and treatment factors, viral properties, or other circumstances

## Introduction

During the first pandemic wave, a substantial decline in mortality was noted among patients hospitalised for COVID-19 in many countries, including Sweden [1-6], but not in all countries [7]. The decline in mortality was believed to be due to a combination of factors, including improved patient triage, decreased patient load, improvements in care and standard medication, and possibly changes in virus virulence [1-4]. Since the relative importance of these factors were unknown, no one could foresee how mortality would change during forthcoming waves.

The aim of the present study was to see how mortality among patients hospitalised for COVID-19 changed during the second pandemic wave.

## Methods

### Study design and setting

Nationwide observational cohort study on SARS-CoV-2-positive individuals treated for COVID-19 in Swedish hospitals, using data compiled by the Swedish National Board of Health and Welfare.

### Participants

The study population was derived from cross-linked national population-based registers, using the same design and statistical methods described previously [1]. All patients hospitalised in Sweden between March 1^st^ and December 31^st^, 2020 and reported to the National Patient Register, with SARS-CoV-2 RNA positivity 14 days before to 5 days after admission, and a discharge code for COVID-19, *i*.*e*. U07.1 or U07.2 according to the *10*^*th*^ *International Statistical Classification of Diseases* (ICD-10), were included.

### Outcome

Study outcome was 60-day mortality, defined as death from any cause within 60 days of index hospital admission. Mortality data (reporting is mandatory by law) were derived from the Swedish Tax Agency.

### Covariate data

Our main exposure of interest was time period of hospital admission. To this end, patients were categorised according to month of admission.

Data on comorbidity (including Charlson Comorbidity Index; CCI), care dependency prior to admission, and ICU data were extracted as described previously [1]. Briefly, to classify comorbidities we used ICD-10 codes registered as in- and outpatient diagnoses during the last five years in the National Patient Register, and prescribed drugs filled out during the year preceding diagnosis, according to the National Prescribed Drug register. Full coding definitions of each comorbidity is given in supplementary methods (Supplementary table S1). The CCI was calculated according to a modified version of the Royal College of Surgeons CCI algorithm (Supplementary table S2). Information on care dependency (nursing home resident or homecare) prior to admission was obtained from the Care and Social Services for the Elderly and for Persons with Impairments Register. In the present study, we have added data on disposable income, education level and main income source, obtained from the Swedish Longitudinal Integrated Database for Health Insurance and Labour Market Studies (LISA database) held by Statistics Sweden.

Information on intensive care treatment, including data on Simplified Acute Physiology Score, version 3 (SAPS3) and oxygenation index (PaO_2_/FiO_2_) (obtained from the initial ICU admission), medication and procedures (obtained from any ICU admission) during ICU care was linked from the Swedish Intensive Care register.

### Statistical analyses

Relative risk (RR) for death within 60 days was estimated using modified Poisson regression models [8] with month of admission as exposure variable of interest. We adjusted the estimates for age (continuous; both a linear and quadratic term), sex (categorical; male/female), CCI (categorical; 0, 1-2, 3-4, 5+), care dependency (categorical; nursing home, homecare, neither), country of birth (categorical; Sweden/other), income quintile (categorical; by quintiles of the distribution in the total population), income source (categorical, categorised as ‘employed/student/caregiver’, ‘disability pension’, ‘sick pay/unemployment’, ‘retirement pension’, ‘financial aid’), and education (categorical; primary school, secondary school, university/college < 3yrs, university/college > 3 yrs), all modelled as main effects. We also performed stratified analysis according to ICU admission during hospital stay. Patients were categorised as “ICU-treated” if they were ever ICU-treated during hospital stay or “non-ICU-treated” if they were never ICU-treated during hospital stay. Estimates for the ICU-treated strata were further adjusted for the Simplified Acute Physiology score, version 3 (SAPS3; linear) from the first ICU episode.

To test for statistical interaction between our main exposure and the covariates, separate models including an interaction term between month of admission and each covariate were constructed. In the interaction model for age, age was entered as a categorical variable (<60, 60-69, 70-79, ≥80 yrs) for increased interpretability. Additionally, when modelling interactions for Income source, all categories except “employed/student/caregiver” and “retired” were collapsed as “other” due to sparse data.

In all regression models, missing data were handled by complete case analysis. Data were complete on all variables except for country of birth (1% missing), CCI (4%), education (6%), income (5%), income source (3%), and SAPS3 (<1% missing).

Duration of hospital stay was defined as number of days between index admission date and last discharge date in cases with sequential admissions in the National Patient Register. A sequential admission was defined as readmission occurring within 1 day.

All data management and statistical analyses were performed using SAS Software SAS Enterprise guide v7.15, SAS Institute Inc., Cary, NC.

### Ethics and Reporting

Ethical approval for the study was obtained from the Swedish Ethics Review Authority, Uppsala (Dnr 2020-04278). The study conforms to the Reporting of Observational Studies in Epidemiology (STROBE) statement.

## Results

### Study population

Altogether, 32 452 patients admitted for COVID-19 to Swedish hospitals March-December 2020 were studied, including 15 839 during the first wave (March-June), 1365 during the inter-wave period (July-September), and 15 248 during the second wave (October-December; figure 1). The peak in number of patients admitted for COVID-19 per month occurred in December, with 8 253 admissions (figure 2A). The proportion of patients admitted to ICU was 14% March-June, 9% July-September, and 10% October-December (figure 2A).

**Figure 1:**
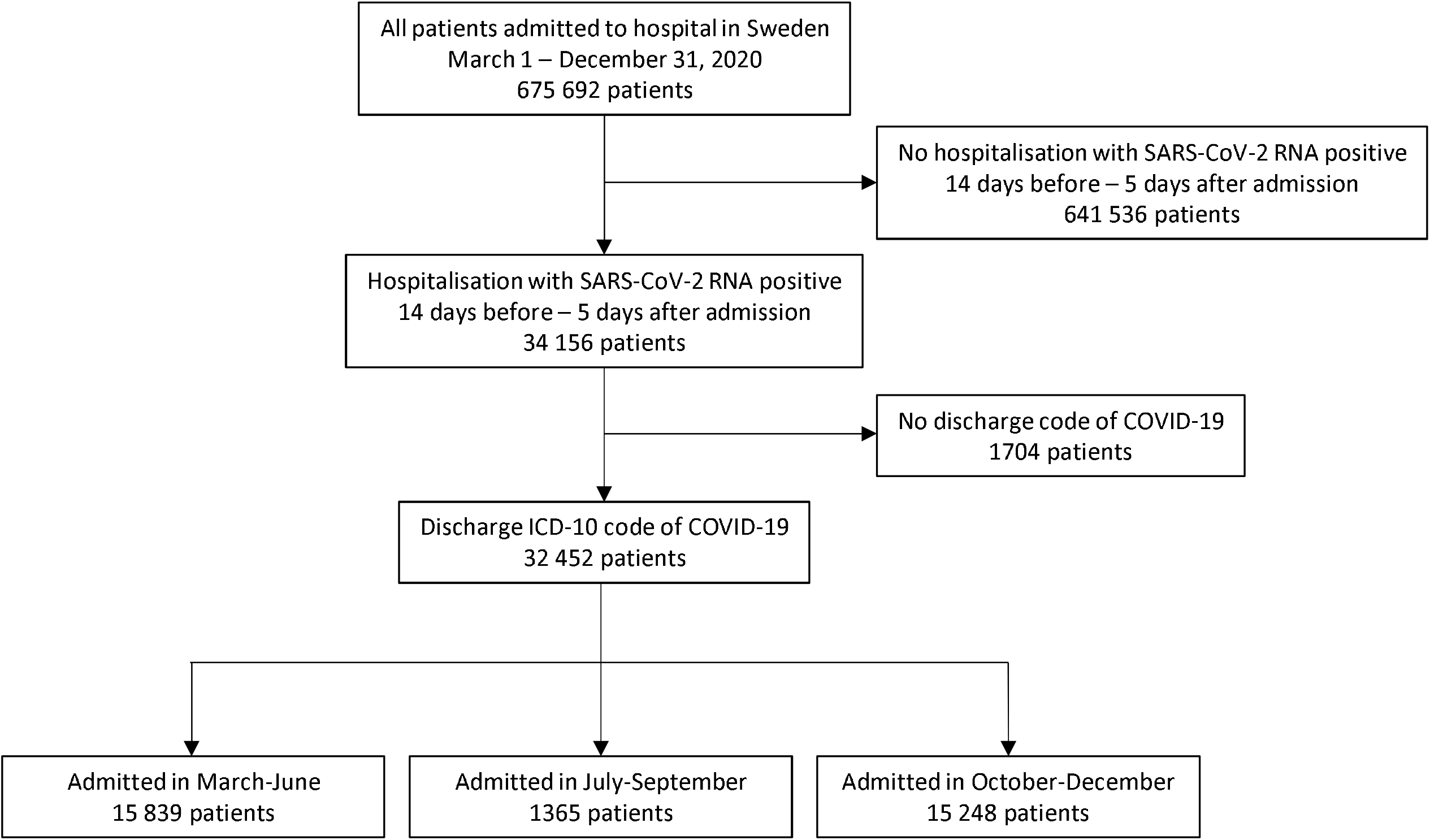
Flowchart of study inclusions and exclusions: Patients hospitalised due to COVID-19 in Sweden March 1 – December 31, 2020.

**Figure 2:**
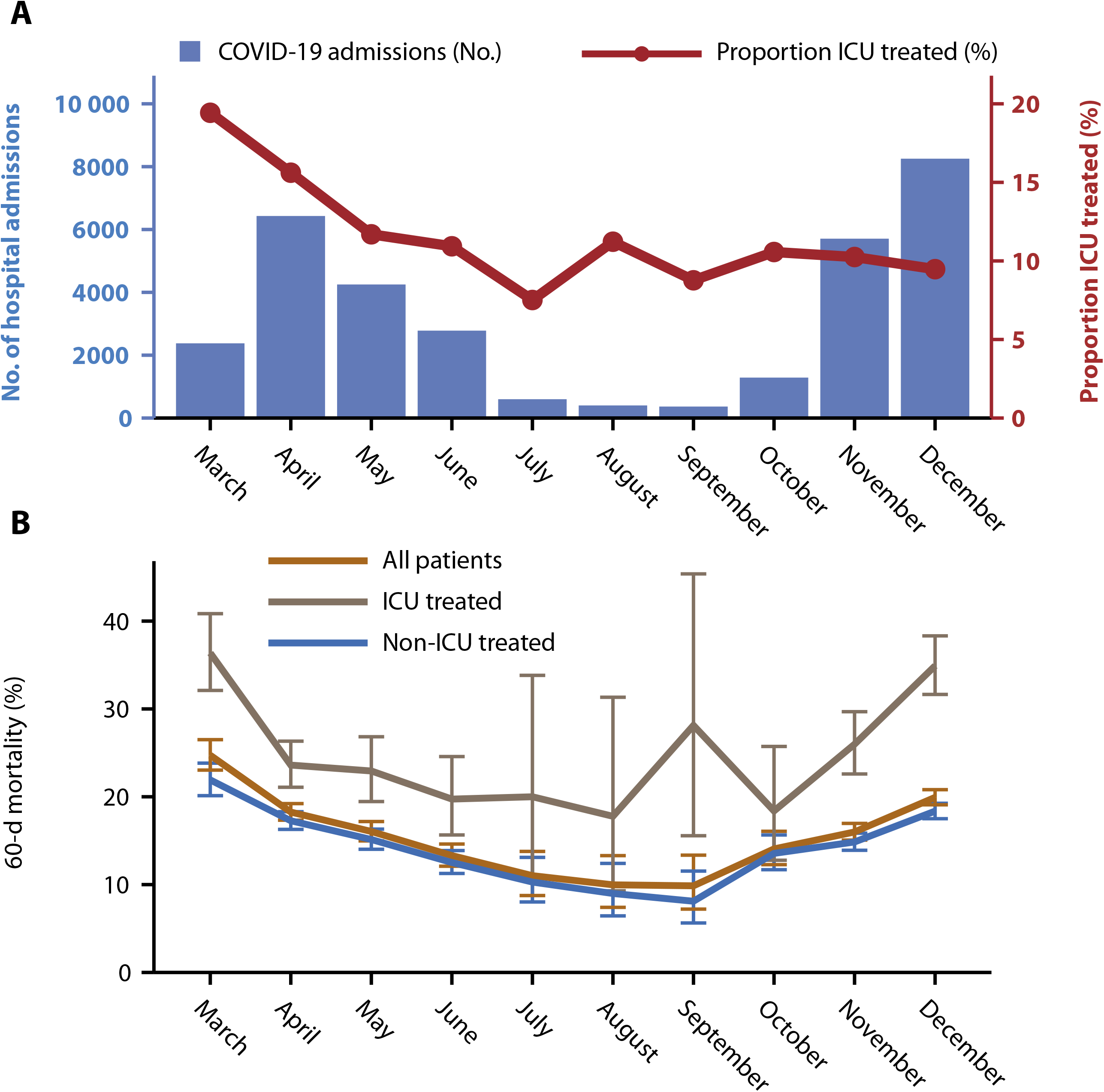
Panel figure, Timeline of the study period: Panel (A), Number of Covid-19 hospital admissions and proportion of intensive care unit (ICU) treatment; Panel (B), Crude 60-day all-cause mortality. Numbers are stratified by month of admission, representing all patients hospitalized for COVID-19 in Sweden March-December 2020.

The median age was 66 years (interquartile age [IQR], 52 to 80 years), and 57% were men. Table 1 shows characteristics of the patients related to month of admission. As noted, between July-September and December, the median age increased from 61 years (IQR 44 to 77 years) to 71 years (IQR 56 to 82 years), the proportion with no comorbidity (CCI 0) decreased from 63% to 58%, the proportion living in nursing homes increased from 5·0% to 8·6%, and the proportion retired increased from 39% to 55%.

**Table 1.**
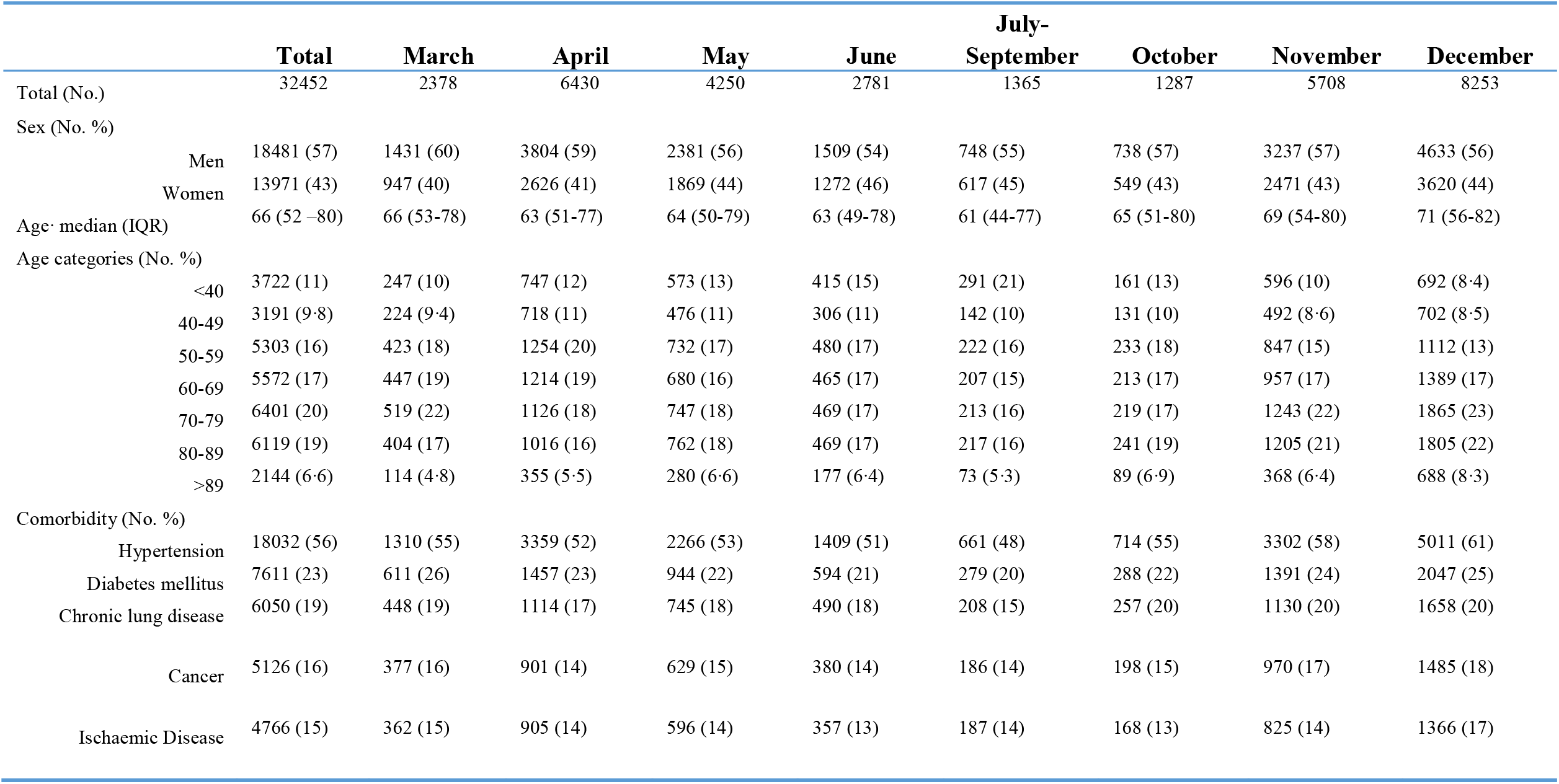
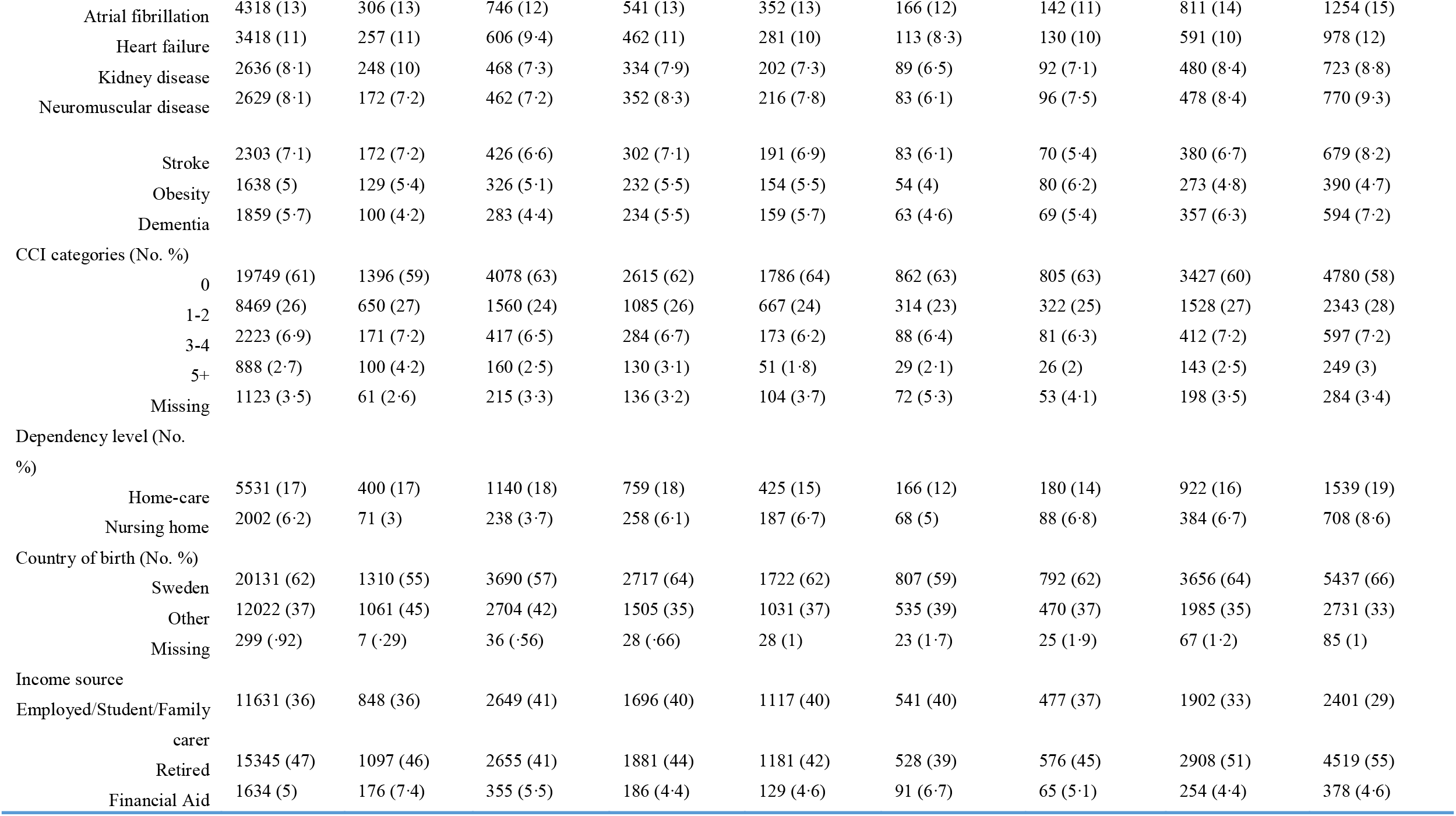
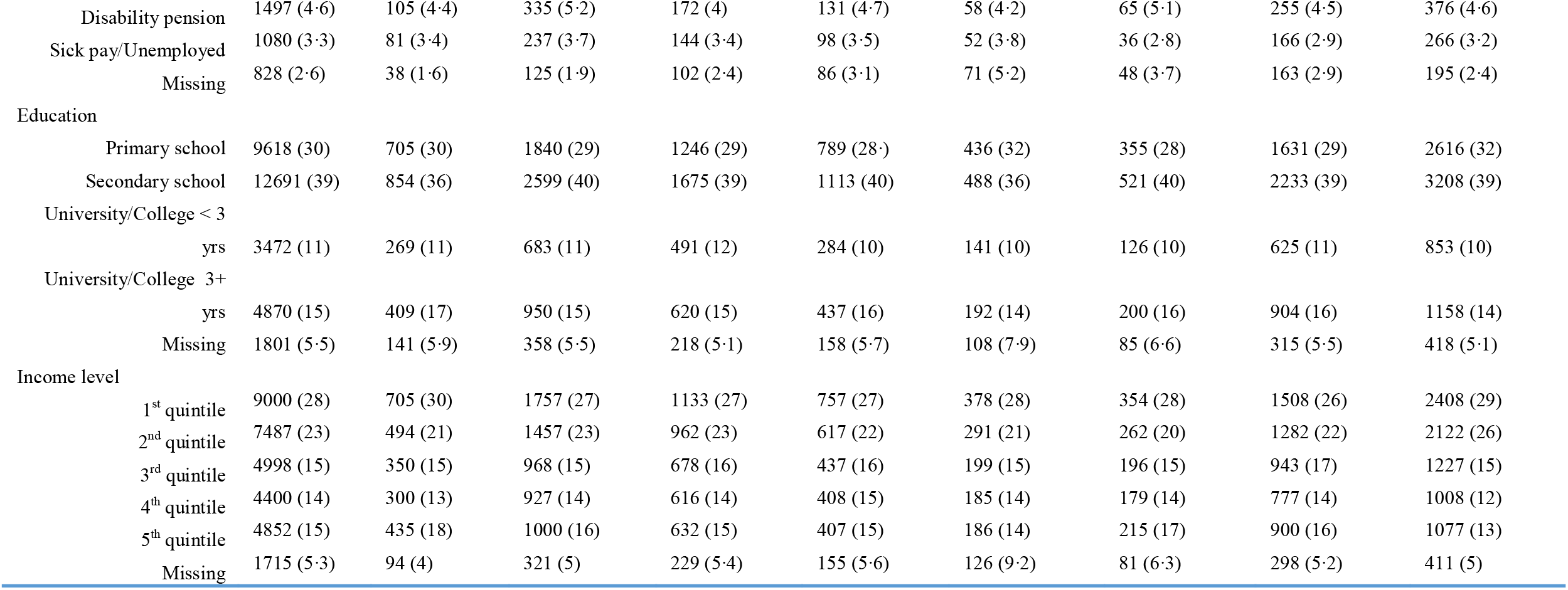
Baseline patient characteristics: total and stratified by month of admission.

Supplementary table S3 shows that the length of hospital stay was substantially longer for ICU-treated patients than for non-ICU-treated patients. Among ICU-treated patients, survivors had longer length of stay than those who died. However, there was not clear difference in length of stay over time.

### Overall mortality

As 5695 patients died within 60 days after admission, overall mortality for March to December 2020 was 17·5% (95% CI, 17·1-18·0); 16·2% (95% CI, 15·8-16·7) for non-ICU-treated patients, and 27·1 % (95% CI, 25·7-28·5) for ICU-treated patients.

A multivariable analysis of RR for death within 60 days is shown in figure 3, where sex, age, income, income source, education level, comorbidity, care dependency, country of birth, and month of admission, are modelled as main effects. Established risk factors such as being male, nursing home resident, and having a CCI of 5+ were all associated with increased mortality.

**Figure 3:**
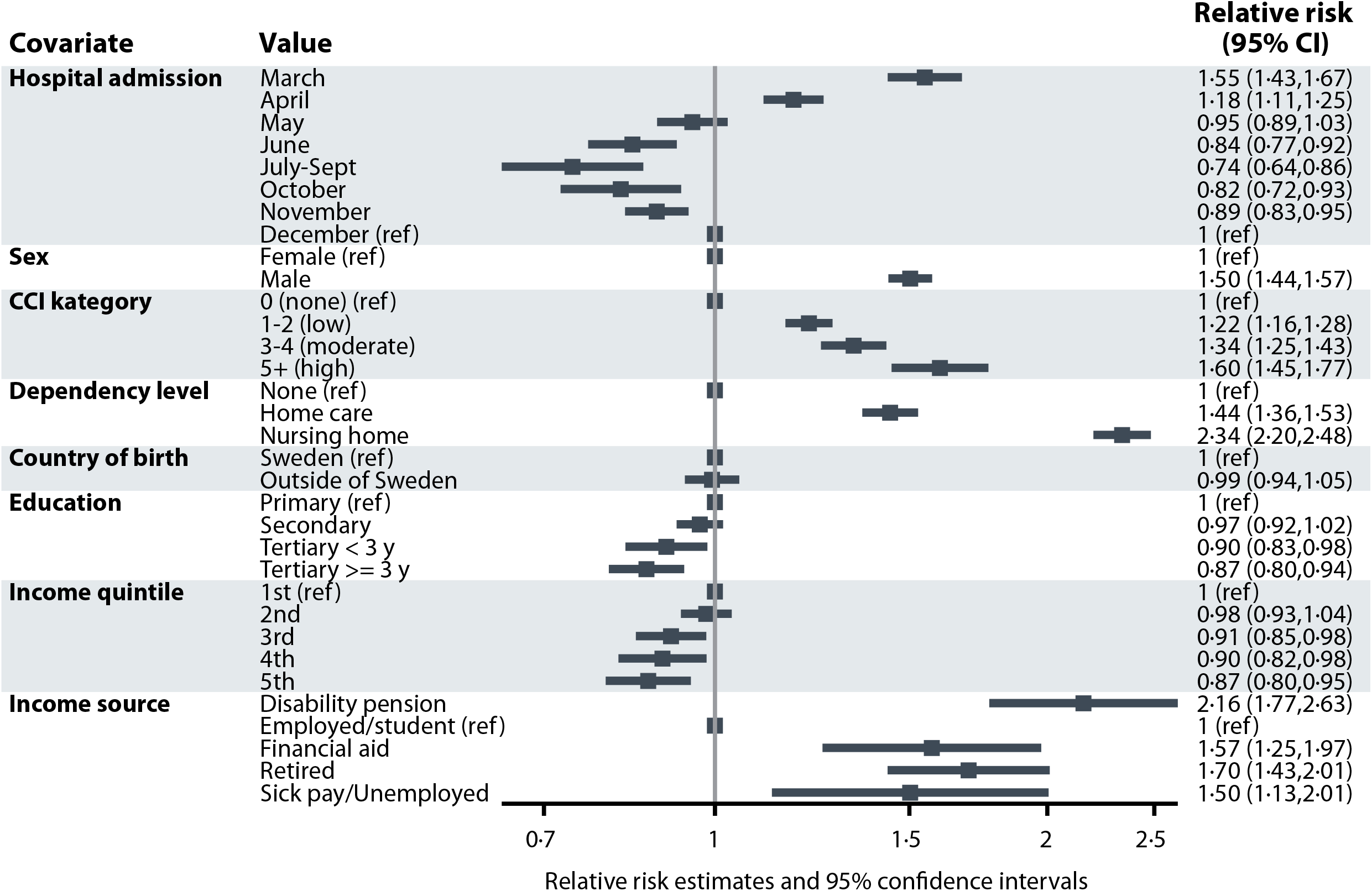
Forest plot: Relative risk of death from any cause within 60 days of hospital admission, for all patients in the cohort. Model adjusted for month of admission, sex, age (continuous, linear and quadratic terms, not shown in the plot), Charlson Comorbidity Index, care dependency, country of birth, education, disposable income quintile and income source.

### Changes in mortality over time

Figure 2B shows that the crude 60-day mortality decreased from 24·7% (95% CI, 23·0%-26·5%) for March to 10·4% (95% CI, 8·9%-12·1%) for July-September (as reported previously [1]), but later increased to 19·9% (95% CI, 19·1 to 20·8) for December. For July to December, the 60-day mortality increased from 10·3% (95% CI, 8·0 to 13·1) to 18·3% (95% CI, 17·5 to 19·3) for non-ICU-treated patients and from 20·0% (95% CI, 11·0 to 33·8) to 34·9% (95% CI, 31·7 to 38·3) for ICU-treated patients.

As can be seen in figure 4, the overall adjusted RR for death within 60 days for December was higher than RRs for June, July-September, October, and November, and was similar to the RR for May. The pattern was similar for non-ICU-treated and ICU-treated patients.

**Figure 4:**
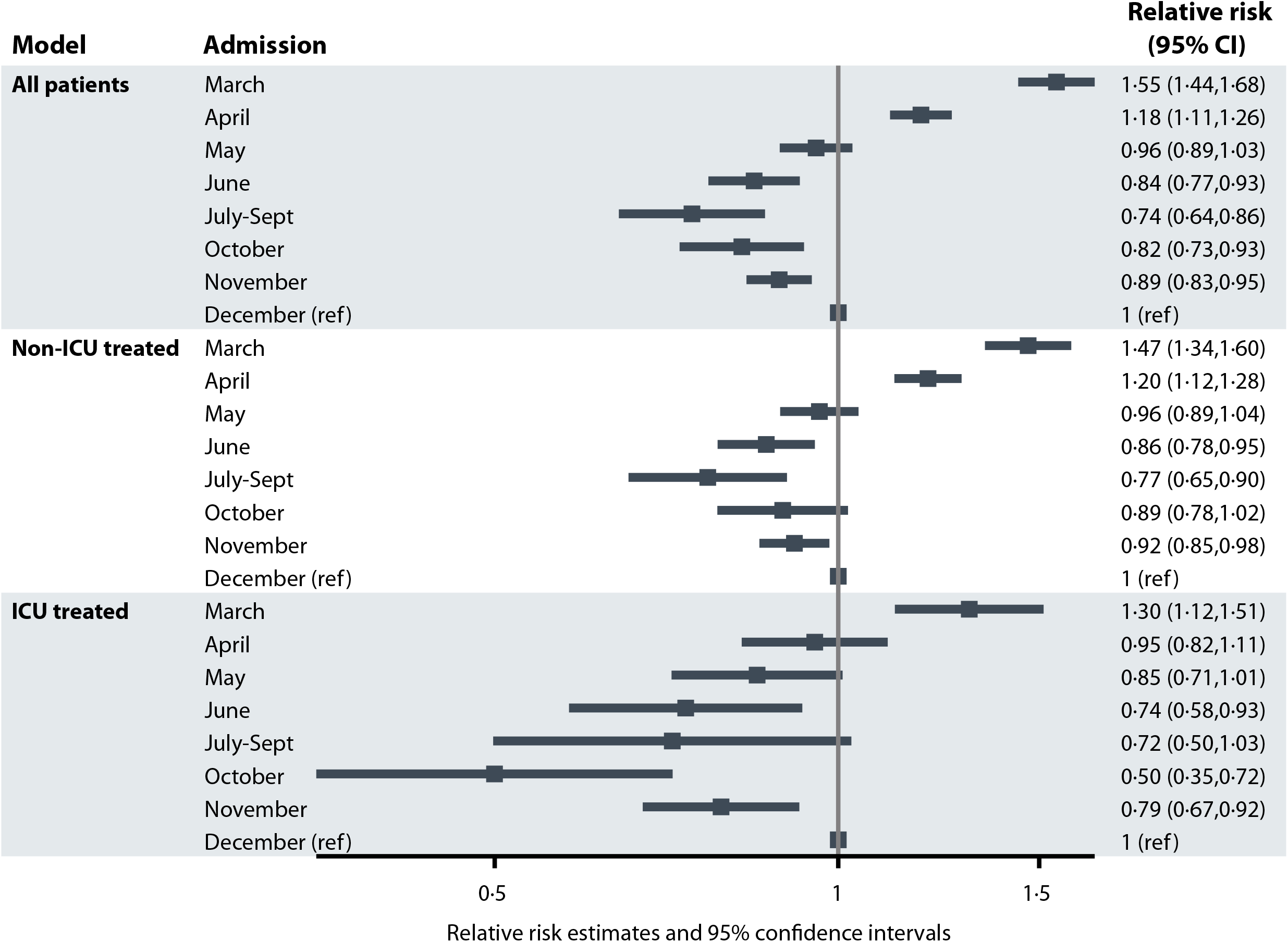
Forest plot: Relative risks with 95% confidence intervals (CI) of death from any cause within 60 days from hospital admission. Upper panel: All hospitalised patients, model adjusted for sex, age (continuous, linear and quadratic terms, not shown in the plot), Charlson Comorbidity Index, care dependency, country of birth, education, disposable income quintile and income source. Middle panel: Patients not treated in ICU, model adjusted for sex, age (continuous, linear and quadratic terms, not shown in the plot), Charlson Comorbidity Index, care dependency, country of birth, education, disposable income quintile and income source. Lower panel: ICU-treated patients, model adjusted for sex, age (continuous, linear and quadratic terms, not shown in the plot), Charlson Comorbidity Index, care dependency, country of birth, education, disposable income quintile, income source and SAPS3 score at first ICU-admission.

Crude mortalities for the different covariates of the study are shown in figure 5. As noted, there was a similar pattern with decreasing mortality for March to July-September and thereafter increasing mortality until December. Figure 6 shows the adjusted RR for death within 60 days for month of admission stratified by each covariate of interest, from the models including both main effect and interaction term. Statistical interaction was found between month of admission and age, comorbidity index, income source, care dependency and country of birth, (p-value for joint test of interaction term < 0·05), but not for sex, disposable income or education (figure 6).

**Figure 5:**
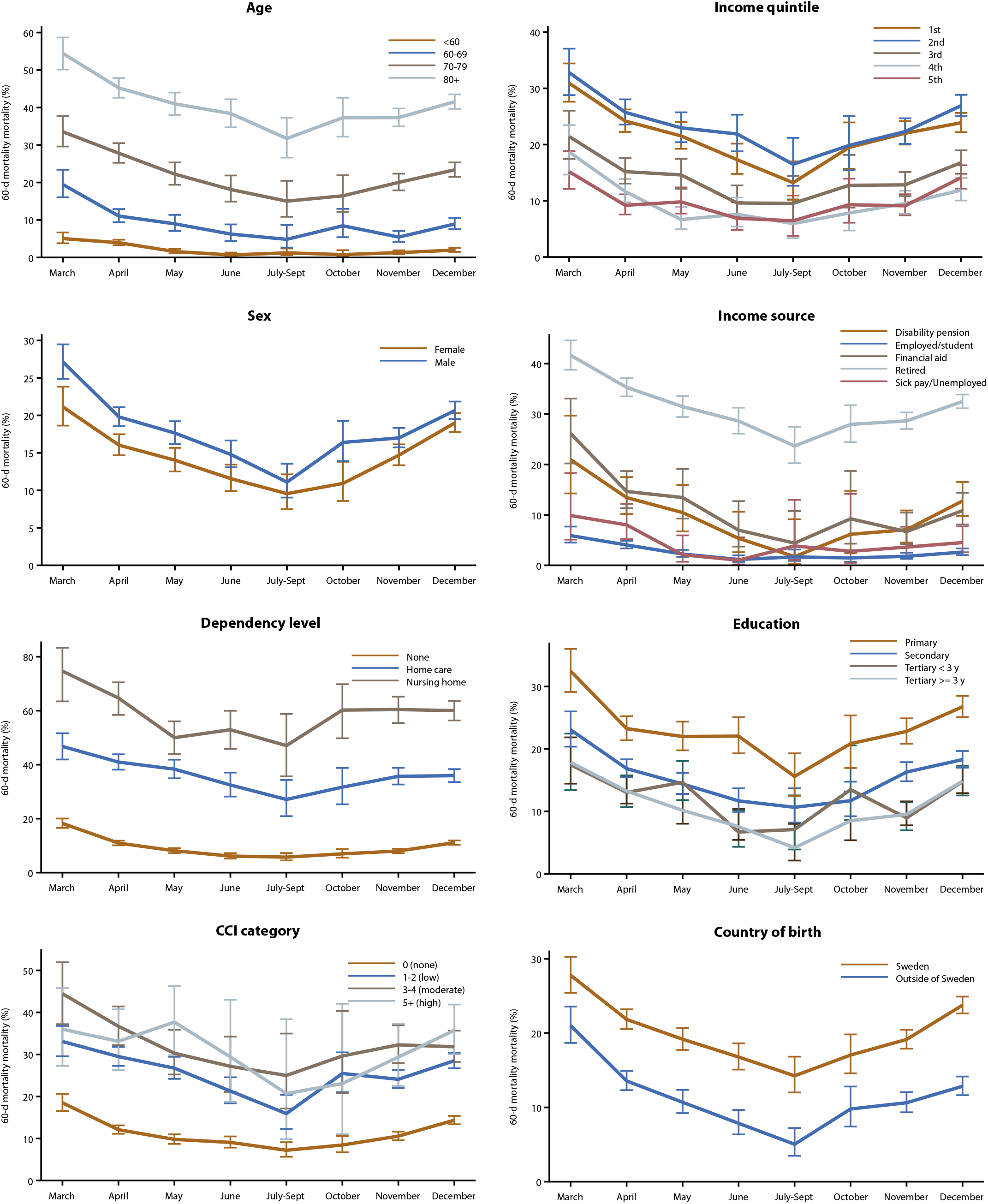
Panel figure: Crude 60-day mortality proportion by (Panel-wise) age, sex, care dependency, Charlson comorbidity index categories, disposable income quintile, income source, education and country of birth. Shown are proportions with 95% confidence intervals per month of hospital admission, separately by categories of each factor.

**Figure 6:**
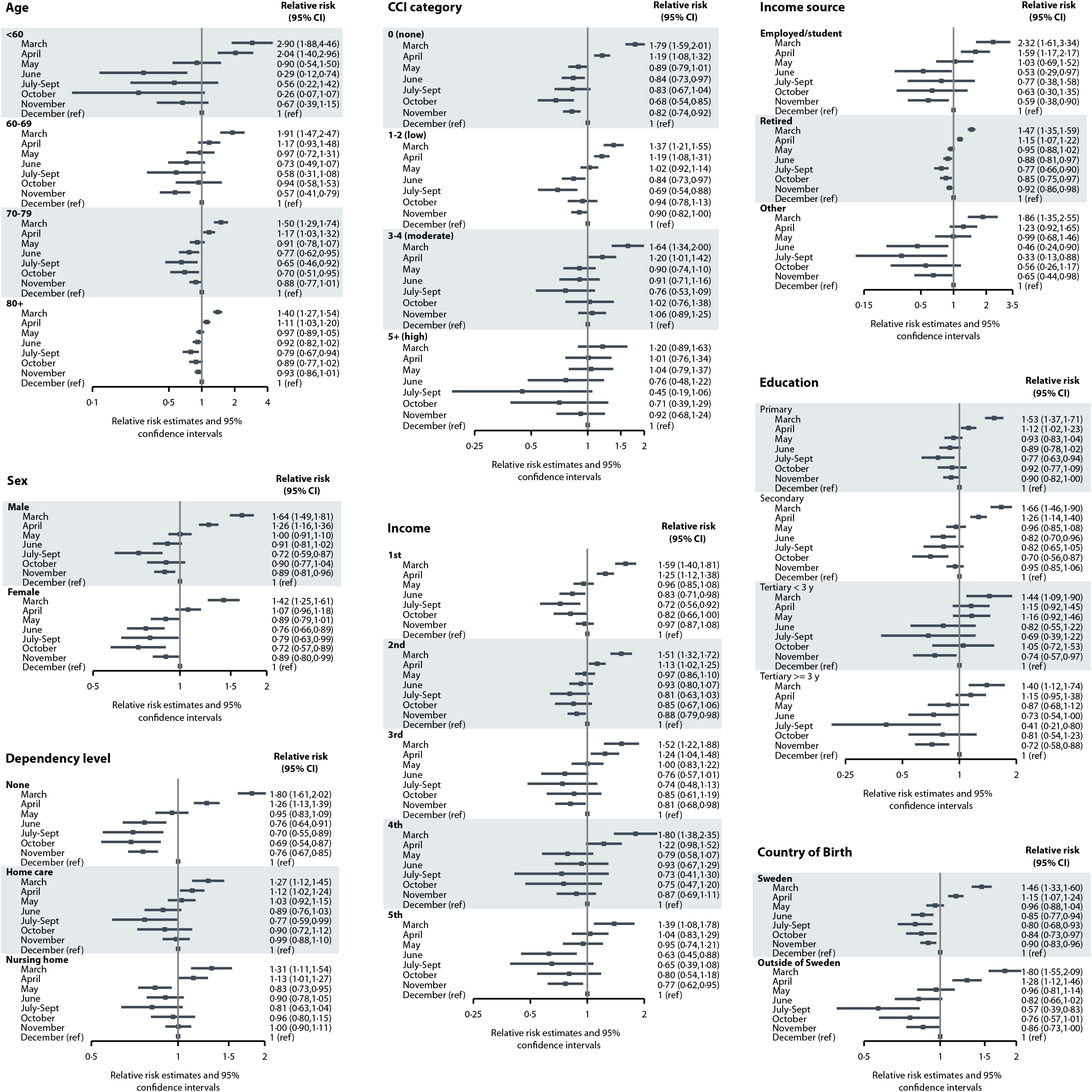
Forest plots showing relative risks (RR) with 95% confidence intervals (CI) of death from any cause within 60 days of hospitalisation, by month of hospital admission, stratified by each level of covariates examined. Estimates were obtained from the models adding an interaction term between month and covariate of interest. One model per covariate of interest underlies the estimates shown, i.e. one for modeling interaction with age, one for modeling interaction with care dependency and so on.

### Changes in characteristics of patients treated on ICU

Altogether 3892 patients were treated in intensive care (table 2) and 2593 (67%) received invasive mechanical ventilation. However, the proportion of ICU patients receiving invasive mechanical ventilation fell over time from 87% in March to 49% in July-September but then increased to 60% in December. It should be noted that the mean PaO_2_/FiO_2_ ratio on admission to ICU gradually decreased from 18 kPa (Standard deviation [SD] 13) in July-September to 15 kPa (SD 8·5) in December. The median hospital stay prior to ICU was 1 day (IQR 1-3 days) for each month March-June, 1 day (IQR 1-2 days) July-September, and 2 days (IQR 0-4) in October, November, and December. It should be noted that corticosteroids were given to <30% of ICU patients March-June, 60% July-September, and >80% October-December.

**Table 2.**
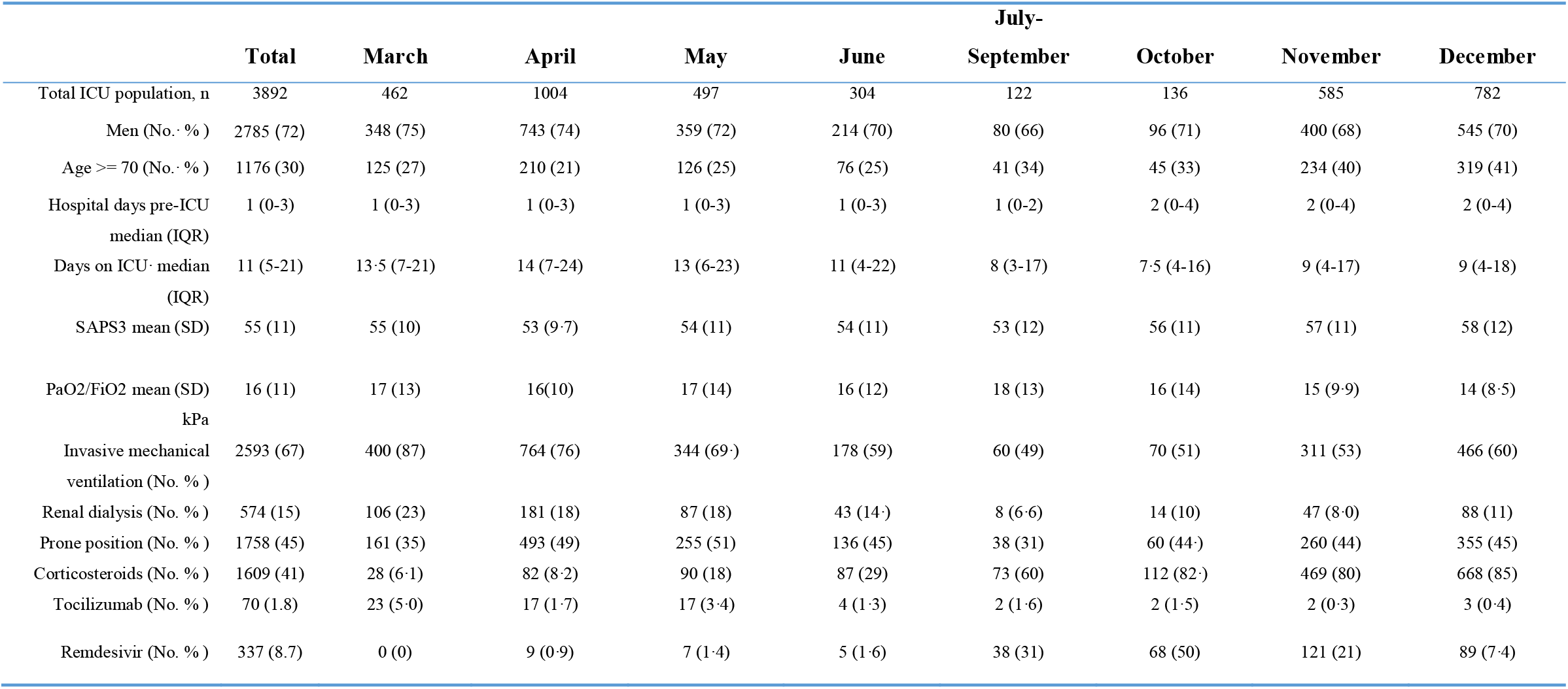
Characteristics of the patient population treated on an intensive care unit (ICU), by month of hospital admission and survival outcome at 60 days follow-up.

## Discussion

The present nationwide study of patients hospitalised for COVID-19 in Sweden showed that the decrease in 60-day mortality after the first pandemic wave turned and increased significantly during the second wave. The crude mortality for December exceeded that for April and the adjusted mortality for December was comparable to that for May.

Although some population-based studies of the second pandemic wave have been published [9-11], very few studies have reported outcome data of hospitalised patients from the second pandemic wave. A nationwide German study [12] showed that ICU admission rate due to COVID-19 during the second wave fell to half that during the first wave of the pandemic. This agrees with our findings (figure 2A). The proportion with invasive mechanical ventilation among ICU treated patients declined in the German study [12], although in the present study it decreased from 87% in March to 49% in July-September, where after it increased to 60% in December (table 2). In the German study, the crude mortality among mechanically ventilated COVID-19 patients was more than 50% throughout 2020. Among Swedish ICU-treated patients, however, crude mortality followed a U-shaped curve, with a maximum of 36% in March declining to 20% during the summer, increasing again to 35% in December (figure 2B).

It is not clear why the mortality among hospitalised patients in Sweden increased during the second wave. It should be noted, however, that population characteristics changed considerably (table 1). Median age increased, the proportion with no comorbidity decreased, the proportion living in a nursing home increased, and the proportion retired increased. Still, even after adjustment for baseline patient characteristics, the increased mortality remained (figure 3, 4) and was seen in all age groups, regardless of comorbidities and care dependency (figure 6).

During the first pandemic wave, management and care of patients with COVID-19 changed considerably [1], and most likely these improvements were partly the reason for the decline in mortality during the first wave. However, during the second half of 2020 no essential changes were made in the national recommendations of standard-of-care for hospitalised COVID-19 patients in Sweden [13], but even so mortality increased.

In studies from the first wave, a high load of hospitalised patients was considered to be associated with a high mortality [7, 14, 15], motivating actions to flatten-the-curve to reduce the pressure on hospitals [16]. The present study with two waves and an inter-wave period supports that there may be an association between number of patients hospitalised for COVID-19 and mortality (figure 2). Among ICU-treated patients in the present study, hospital stay prior to ICU admission was one day (IQR 0-2) July-September, and two days (IQR 0-4) October-December, and the mean PaO_2_/FiO_2_ ratio on ICU admission gradually decreased from July-September to December (table 2). This could indicate that ICU admission was delayed in the second wave. It was shown prior to the pandemic that hospital capacity strain is associated with increased mortality [17].

Another reason for the relationship between high mortality and peak in admission rate could be high exposure to circulating virus in the community, leading to heavier viral inoculates, perhaps causing more severe disease [18].

It is also possible that the increase in mortality during the second half of 2020 was caused by mutations of the SARS-CoV-2 virus. The SARS-CoV-2 virus variant of concern (B.1.1.7), which was first detected in the United Kingdom has been associated with an enhanced mortality among out-patients with SARS-CoV-2 infection [19, 20]. However, as virus variants of concern occurred only sporadically in Sweden before January 2021 [21], they were most likely not involved in the increased second wave mortality that was noted in this study.

The present study has a number of strengths. First, it is a nationwide study covering all hospitals in Sweden, which reduced the risk of selection bias. The unique national personal identification number of all Swedish residents enabled cross-linkage of registers at the individual level. Second, the use of 60-day mortality provided a robust outcome measure with a follow-up long enough to enable hospital discharge or death of most patients. Third, the study results cannot have been influenced by anti-SARS-CoV-2 vaccination, since vaccination in Sweden began December 27^th^, 2020. Forth, the study results were probably not influenced by virus variants of concern, as discussed above [21].

The study has limitations as mentioned before [1]. First, data on disease severity of non-ICU-treated patients and data on do-not-resuscitate orders were not available. Second, we did not have access to data on drug treatment of non-ICU-treated patients and could not assess its impact on outcome. Third, the study lacked information on hospital bed occupancy and staff-to-patient ratio, which is important when assessing association between patient load and outcome.

In conclusion, the decrease in mortality that was noted during the summer of 2020 did not continue during the second pandemic wave. Instead the 60-day mortality increased. Focused research is urgent to describe if this increase was caused by a high load of patients, management and treatment, viral properties, or other factors.

## Data Availability

Data sharing
The data underlying this article cannot be shared publicly due to regulations under Swedish law. According to the Swedish Ethics Review Act, the General Data Protection Regulation, the Public Access to Information and Secrecy Act, data can only be made available, after legal review, for researchers who meet the criteria for access to this type of confidential data. Requests regarding data in this paper may be made to the senior author.

## Contributors

All authors conceived and designed the study.

KS, EW, SW, ABB, MH, JH and HH acquired data.

EW and JH performed analyses and interpreted these together with KS,

SW and HH.

EW and JH verified the underlying data in the article.

EW and JH drafted and finalised all tables and figures.

KS and EW contributed equally.

KS drafted the first version of the manuscript.

All authors had full access to data in the study and accept responsibility for submission for publication.

All authors revised the manuscript critically for important intellectual content and approved the final version for submission.

## Acknowledgments

This study was funded by Sweden’s National Board of Health and Welfare.

## Data sharing

The data underlying this article cannot be shared publicly due to regulations under Swedish law. According to the Swedish Ethics Review Act, the General Data Protection Regulation, the Public Access to Information and Secrecy Act, data can only be made available, after legal review, for researchers who meet the criteria for access to this type of confidential data. Requests regarding data in this paper may be made to the senior author.

