## Supplementary for "Second wave mortality among patients hospitalised for COVID-19 in Sweden: a nationwide observational cohort study"

### Supplementary material

**Supplementary table S1.** Definition of comorbidities using ICD-10 and ATC-codes. A comorbidity was considered to be present if: 1. At least one of the ICD-codes were registered during the period 2015-2020, excluding 30 days before admission in the Inpatient or Outpatient Register; and/or 2. At least one prescription for a drug with at least one of the ATC-codes below were expedited during the period 2019-2020, excluding 30 days before admission, according to the Prescribed Drug Register

| | ICD-10 | ATC |
|---|---|---|
| Ischaemic disease | I20 I21 I22 I23 I24 I25 | N02BA C01DA B01AC24 |
| Heart Failure | I110 I130 I132 I50 | |
| Hypertension | I109 I11 I12 I13 I15 | C02 (exkl C02AC02) C03 C09 C08CA C07AB02 |
| Atrial fibrillation | I48 | |
| Stroke | I60 I61 I62 I63 I64 I69 | |
| Diabetes mellitus | E10 E11 E12 E13 E14 | A10 |
| Chronic lung disease | J41 J42 J43 J44 J45 J46 J47 | R03AK R03AL R03BA R03AC12 R03AC13 R03AC18 R03AC19 R03CC12 R03BB04 R03BB05 R03BB06 R03BB07 |
| Dementia | F00 F01 F02 F039 G30 F107A | N06D |
| Cancer | C00-C97 Z85 | |
| Kidney disease | I12 I13 N00 N01 N02 N03 N04 N05 N07 N08 N11 N14 N18 N19 E102 E112 | |
| Neuromuscular disorders | G10 G11 G12 G13 G14 G20 G21 G22 G23 G24 G25 G26 G30 G31 G32 G35 G36 G37 G70 G71 G72 G73 G80 G81 G82 G83 | |
| Obesity | E66 | A08 |

**Supplementary table S2**. ICD-10 codes and weights defining comorbidity categories for calculating the Charlson Comorbidity Index. Modified from J. N. Armitage and J. H. van der Meulen: “Identifying comorbidity in surgical patients using administrative data with the Royal College of Surgeons Charlson Score”. British Journal of Surgery 2010; 97: 772–781. The index was calculated based on diagnoses reported to the National Patient Register 2015 - 2019

| Category | Weight | ICD-10 |
|---|---|---|
| Myocardial infarction | 1 | I252 |
| Acute myocardial infarction | 1 | I21 I22 I23 |
| Congestive heart failure | 1 | I11 I13 I255 I42 I43 I50 I517 |
| Peripheral vascular disease | 1 | I70 I71 I72 I73 I770 I771 K558 K559 Z958 Z959 K551 R02 |
| Cerebrovascular disease | 1 | G45 G46 I6 |
| Dementia | 1 | F00 F01 F02 F03 G30 G31 A810 F051 |
| Chronic pulmonary disease | 1 | I26 I27 J40 J41 J42 J43 J44 J45 J47 J60 J61 J62 J63 J64 J65 J66 J67 J684 J701 J703 |
| Acute chronic pulmonary disease | 1 | J46 |
| Rheumatic disease | 1 | M05 M06 M32 M33 M34 M35 M36 M09 M120 M315 |
| Liver disease | 1 | K70 K71 B18 I85 I864 I982 K721 K729 K76 R162 Z944 |
| Diabetes mellitus without complications | 1 | E100 E1032 106 E108 E109 E110 E111 E116 E118 E119 E120 E121 E126 E128 E129 E130 E131 E136 E138 E139 E140 D141 E146 E148 E149 |
| Diabetes mellitus with complications | 2 | E102 E103 E104 E105 E107 E112 E113 E114 E115 E117 E122 E123 E124 E125 E127 E132 E133 E134 E135 E137 E142 E143 E144 E145 E147 |
| Hemiplegia/Paraplegia | 2 | G81 G83 G114 |
| Renal disease | 2 | I12 I13 N01 N03 N05 N07 N08 N18 N25 Z49 Z940 Z992 |
| Acute renal disease | 2 | N171 N172 N19 |
| Malignancy | 2 | C0 C1 C20 C21 C22 C23 C24 C25 C26 C30 C31 C32 C33 C34 C37 C38 C39 C40 C41 C45 C46 C47 C48 C49 C50 C51 C52 C53 C54 C55 C56 C57 C58 C6 C70 C71 |

| | | |
|---|---|---|
| | | C72 C73 C74 C75 C76 C80 C81 C82 C83 C84 C85 C90 C91 C92 C93 C94 C95 C96 C97 C43 C88 |
| Metastatic tumours | 6 | C77 C78 C79 |
| AIDS/HIV | 6 | B20 B21 B22 B23 B24 |

**Supplementary table S3**. Duration of hospital stay in days, by period of admission, discharge status and patient category. Shown are number of patients, mean and median hospital stay in days with interquartile range (IQR; 25th and 75th percentiles).

| Patient category | Discharge status | Admission period | Number of patients | Hospital duration (days) | | | |
|---|---|---|---|---|---|---|---|
| | | | | Mean | Median | 25th Pctl | 75th Pctl |
| **Non- ICU treated** | **Alive** | March | 1548 | 6,8 | 5 | 2 | 9 |
| | | April | 4619 | 7,1 | 5 | 2 | 9 |
| | | May | 3284 | 7,2 | 5 | 2 | 9 |
| | | June | 2228 | 6,6 | 4 | 2 | 9 |
| | | July-Sept | 1166 | 6,0 | 4 | 2 | 7 |
| | | October | 1043 | 6,7 | 5 | 2 | 8 |
| | | November | 4543 | 6,9 | 5 | 2 | 9 |
| | | December | 6420 | 7,5 | 5 | 2 | 10 |
| | **Deceased** | March | 367 | 7,1 | 6 | 4 | 9 |
| | | April | 805 | 7,1 | 6 | 3 | 9 |
| | | May | 463 | 7,5 | 6 | 4 | 10 |
| | | June | 248 | 7,8 | 6 | 4 | 10,5 |
| | | July-Sept | 76 | 8,4 | 6,5 | 3 | 12 |
| | | October | 108 | 10,1 | 8 | 4 | 12 |
| | | November | 580 | 9,2 | 7 | 4 | 12 |
| | | December | 1044 | 8,6 | 7 | 4 | 12 |
| **ICU treated** | **Alive** | March | 293 | 36,7 | 28 | 17 | 48 |
| | | April | 761 | 36,6 | 27 | 16 | 49 |
| | | May | 379 | 39,5 | 27 | 16 | 50 |
| | | June | 240 | 34,3 | 20,5 | 12 | 46,5 |

| | | | | | | |
|---|---|---|---|---|---|---|
| | July-Sept | 98 | 27,9 | 15,5 | 8 | 34 |
| | October | 108 | 28,8 | 21 | 12 | 41 |
| | November | 414 | 28,1 | 20 | 12 | 38 |
| | December | 450 | 27,1 | 21 | 12 | 37 |
| **Deceased** | March | 168 | 15,4 | 12 | 7 | 20,5 |
| | April | 243 | 20,4 | 15 | 9 | 25 |
| | May | 116 | 17,8 | 14 | 7 | 24 |
| | June | 63 | 21,4 | 18 | 7 | 31 |
| | July-Sept | 24 | 20,5 | 19 | 12,5 | 24,5 |
| | October | 26 | 20,2 | 15,5 | 8 | 24 |
| | November | 156 | 22,5 | 19 | 12 | 27 |
| | December | 266 | 20,2 | 19 | 10 | 27 |
